# Predicting Early-Onset Colorectal Cancer in Individuals Below Screening Age Using Machine Learning and Real-World Data

**DOI:** 10.1101/2024.07.17.24310573

**Authors:** Chengkun Sun, Erin M. Mobley, Michael B. Quillen, Max Parker, Meghan Daly, Rui Wang, Isabela Visintin, Ziad Awad, Jennifer Fishe, Alexander Parker, Thomas J. George, Jiang Bian, Jie Xu

## Abstract

**Background:** Colorectal cancer (CRC) is now the leading cause of cancer-related deaths among young Americans. Our study aims to predict early-onset CRC (EOCRC) using machine learning (ML) and structured electronic health record (EHR) data for individuals under the screening age of 45.

**Methods:** We identified a cohort of patients under 45 from the OneFlorida+ Clinical Research Consortium. Given the distinct pathology of colon cancer (CC) and rectal cancer (RC), we created separate prediction models for each cancer type with various ML algorithms. We assessed multiple prediction time windows (0, 1, 3, and 5 years) and ensured robustness through propensity score matching (PSM) to account for confounding variables. Model performance was assessed using established metrics. Additionally, we employed the Shapley Additive exPlanations (SHAP) to identify risk factors for EOCRC.

**Results:** Our study yielded results, with Area Under the Curve (AUC) scores of 0.811, 0.748, 0.689, and 0.686 for CC prediction, and 0.829, 0.771, 0.727, and 0.721 for RC prediction at 0, 1, 3, and 5 years, respectively. Notably, predictors included immune and digestive system disorders, along with secondary cancers and underweight, prevalent in both CC and RC groups. Blood diseases emerged as prominent indicators of CC.

**Conclusion:** This study highlights the potential of ML techniques in leveraging EHR data to predict EOCRC, offering valuable insights for potential early diagnosis in patients who are below the recommended screening age.

## 1. Introduction

Colorectal cancer (CRC) is a significant public health challenge, ranking as the third leading cause of cancer-related mortality among both males and females in the United States.^1^ It is estimated that in 2023, approximately 153,020 individuals will be diagnosed with CRC, and 52,550 will succumb to the disease.^1^ While cancer is typically a disease of older age, a concerning trend has emerged – the increasing incidence of early-onset colorectal cancer (EOCRC) in individuals younger than the age of 50 years.^1,2^ This increased incidence has led the US Preventive Services Task Force to modify its recommendations, lowering the age to start CRC screening to age 45.^3^ Patients diagnosed with EOCRC tend to present at later stages and face lower disease-specific survival rates, underscoring the need for early detection and treatment initiation.^4^ Nevertheless, challenges in addressing EOCRC are compounded by poorly defined risk factors and the role of diagnostic delays. As a result, early prediction and comprehensive understanding of the risk factors of EOCRC are essential for prevention and treatment, particularly for patients who fall below the recommended screening age.

The rapid integration of artificial intelligence (AI) and big data analytics has significantly expanded the horizons of medical research and clinical care.^5^ Diverse data sources, including imaging and genomic data, have been harnessed for CRC detection through the application of statistical and machine learning (ML) algorithms. Some approaches have included the analysis of tumor DNA and circulating RNA expression profiling data to identify potential pathogenic factors.^6,7^ Additionally, computer tomography (CT)—based radionics, combined with ML algorithms, have been employed to predict the KRAS mutation in CRC patients, demonstrating the potential of ML in clinical decision support.^8^ Further, a random forest (RF) model trained with standard clinical and pathological prognostic variables, coupled with MRI images, achieved an impressive Area Under the Curve (AUC) score of 0.94 when predicting survival in CRC patients, highlighting the importance of MRI-based texture features patient survival prediction.^9^ However, imaging data produces a small number of unexplainable predictors (∼100), and does not consistently improve diagnostic accuracy and disease prediction, especially when only using imaging data.^10^ Furthermore, advanced imaging modalities and genomic data can be costly, with limited accessibility, and lack diversity and representativeness in samples, which could impact timely and accurate diagnosis for all individuals affected by EOCRC or widen already present disparities in patient outcomes.

In contrast to imaging and genomic data, structured data from the electronic health record (EHR) offer a more accessible and cost-effective data source for initial research. Originally designed for administrative and billing purposes, structured EHR data have evolved into valuable tools for healthcare research, capturing a wealth of patient information, including clinical diagnoses, procedures, medications, and laboratory results, among others.^11^ The integration of ML and deep learning with EHR data has demonstrated substantial potential for disease prediction, including Alzheimer’s disease, gestational diabetes mellitus (GDM), and coronary heart disease (CHD).^12–14^ In the context of CRC, several ML approaches have been employed to predict the risk of the disease. For example, Shanbehzadeh et al. used structured EHR data and four data mining algorithms to predict CRC risk, identifying critical attributes for the prediction model using the weight statistical Chi-square test.^15^ However, the weight statistical Chi-square test assumes independence among variables, which may not hold true in complex datasets where variables are likely correlated. Another study leveraged convolutional neural networks to predict CRC risk based on the structured EHR data from the Taiwan National Health Insurance database.^16^ Hisham et al. explored multiple ML methods to construct predictive models for CRC among patients aged between 35 and 50.^17^ However, these studies faced challenges in effectively matching cases and control groups, leading to increased bias and concerns regarding confounding. Furthermore, another limitation across studies is the failure to distinguish between colon and rectal cancers, despite the differences in clinical presentation, molecular carcinogenesis, pathology, surgical topography and procedures, and multimodal treatment strategies between these two cancers.^18^ Additionally, the lack of model explanations regarding clinical diagnosis of CRC undermined the interpretability and reliability of their strategies. As a result, there is a pressing need for improved methodologies to enhance the reliability and understanding of ML models in EOCRC prediction. In light of these gaps in existing literature, our primary objective is to build separate ML models for the prediction of colon and rectal cancers in patients prior to reaching the screening age of 45 years, leveraging EHR data to identify potential unique risk factors for each cancer type. To achieve this goal, we employed a range of traditional ML models to predict these cancers at various time intervals before their onset (following the setting of Li et al^12^). To mitigate potential data bias and confounding issues, we implemented propensity score matching (PSM) to establish a comparable matched control group.^19^ Additionally, we utilized the Shapley Additive exPlanations (SHAP)^20^ for model interpretation, thereby enhancing our ability to discern the contribution of individual features. By improving interpretability, our aim is to pinpoint the risk factors that pre-date the development of EOCRC.^21^

## 2. Materials and Methods

### 2.1. Data source and study population

This study used de-identified EHR data from the OneFlorida+ Clinical Research Consortium funded by the Patient-Centered Outcomes Research Institute (PCORI), as one of the 8 clinical data research network contributing to the National Patient-Centered Clinical Research Network (PCORnet).^22^ The OneFlorida+ data encompasses a wide range of patient characteristics from health systems across the southeast, including EHR data collected using the PCORnet Common Data Model^22^ regarding demographics, diagnoses, medications, procedures, vital signs, lab tests, and more.

The construction of our study cohort using OneFlorida+ is outlined in Figure 1. OneFlorida+ identified individuals from the OneFlorida+ network, with encounters from January 2012 to January 2023 who met our inclusion criteria as either a case or control. We identified cases of colon cancer (CC) using the International Classification of Diseases (ICD)-9 code of C18* or C49A4 or ICD-10 code of 153*, or rectal cancer (RC) cases with the ICD-9 code of C19*, C20*, C21.0, C21.1, and ICD-10 code of 154.0 and 154.1. The initial cohort consisted of 68,293 CRC cases (54,939 CC cases, 29,592 RC cases), and 589,823 controls. From those, we excluded patients diagnosed with both CC and RC, other prior cancers, or who were diagnosed ≥45 years of age. Our final study cohort comprised 1,358 CC cases with 25,485 controls and 560 RC cases with 22,648 controls.

**Figure 1.**
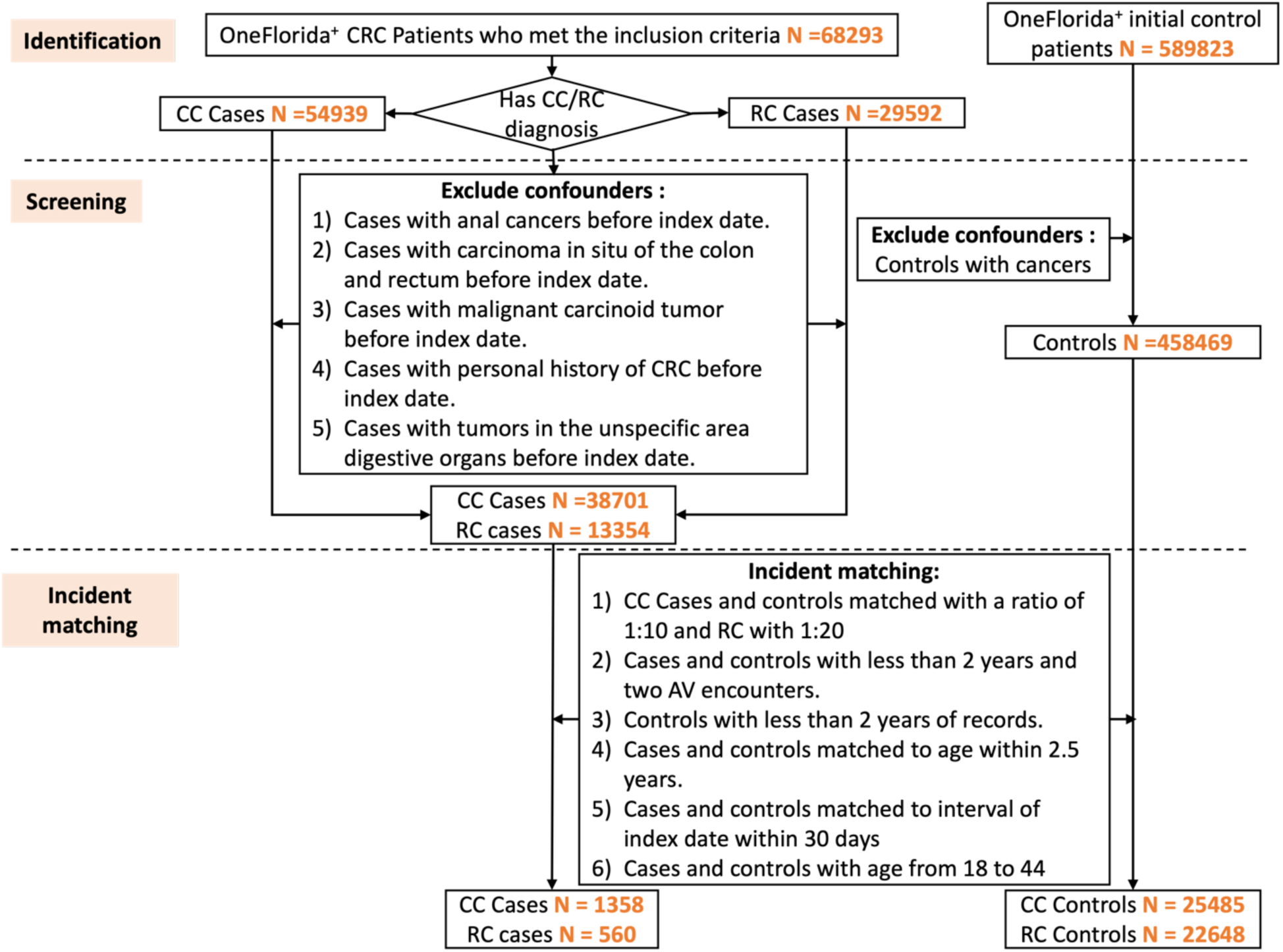
Flowchart of patient selection from OneFlorida+.

**Figure 2.**
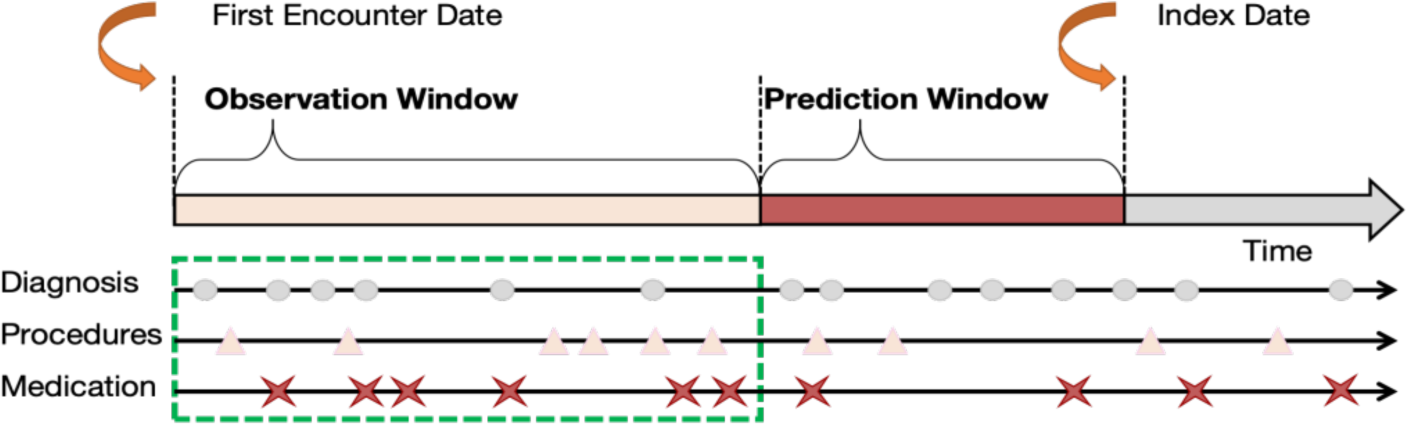
Visualization of the observation and prediction windows for the prediction task. The index date for CRC cases is the date of diagnosis. For the control group, the index date is defined as the closest encounter date to the diagnosis date of the matched case group. The prediction window is the time period before the index date during which CRC cases are predicted. The observation window refers to the specific period during which data is collected or observed for analysis.

We used an incident matching process to match cases and controls to ensure a fair comparison across these groups. Initially, we retained cases and controls with more than two years of records and at least two encounters before the first onset date of either colon or rectal cancer and ensured that the age gap between matched cases and controls was within 2.5 years. By calculating propensity scores based on race, ethnicity, sex, and birth year (within 2.5 years), we employed a narrow caliper of 0.05 with a nearest neighbor approach to achieve a 1:5 case-to-control ratio for each prediction window group.^23^ This rigorous methodology ensures a balanced study population for reliable analysis and EOCRC prediction.

### 2.2. Study setting

Further, we incorporated a range of different observation periods and prediction windows to test our prediction algorithms, considering the different use cases. We considered four different prediction windows: 0-year, 1-year, 3-year, and 5-year before CRC diagnosis.

### 2.3. Data preprocessing

The predictors we extracted include demographics, vitals, diagnoses, medications, and procedures documented throughout the observation periods. Age at index date was calculated and categorized into three groups (e.g., 18-29, 30-39, 40-44). One-hot encoding24 was used to represent age groups, race, and sex variables. For missing data, we imputed the missing values with the mean of the numerical data derived from the entire sample within each prediction window group. Furthermore, BMI data was categorized into clinically relevant groups, including underweight (≤18.5), normal (18.5-23), overweight (23-30), and obese (≥30). Diastolic and systolic measurements were categorized into distinct hypertension stages.

Diagnoses, which were initially represented using ICD-9 and ICD-10 codes, were subjected to a data dimensionality reduction process that mapped them into Phecodes.^25,26^ Revenue codes and Current Procedural Terminology (CPT) codes^27^ were leveraged to capture billed medical procedures. To integrate these data, we also employed the Clinical Classifications Software (CCS) code.^28^ For drug information, National Drug Code (NDC)^29^ and RxNorm codes were used for encoding. NDC codes were mapped into RxNorm codes, and further consolidated into Anatomical Therapeutic Chemical (ATC) classes.^30^ To ensure completeness, all features that could not be mapped were retained to prevent any missing information. These steps to transform the data enhanced interpretability and relevance of our predictive models.

### 2.4. Experiments and validation

We explored several widely used ML models, including linear models such as logistic regression (LR) and the support vector machine (SVM), as well as nonlinear models like XGBoost and RF. We adopted two modeling strategies, including (1) prediction without CRC-related features; and (2) prediction without cancer-related features, covering the CRC-related features. For the first strategy, features that may be indicative of CRC differential diagnoses (e.g., neoplasm of unspecified nature of digestive system) or treatments for CRC (e.g., chemotherapy, radiotherapy) were removed from the models and not used as predictors. For the second strategy, we took a more stringent approach by eliminating all diagnoses, drugs, and procedures that could be associated with any cancer from the extracted predictors. This step aimed to identify risk factors while eliminating the influence of other types of cancers, enabling us to focus exclusively on non-cancer-related predictors. Regardless of the feature engineering strategy, we maintained a consistent experimental setup.

The entire dataset was randomly split into a training dataset and a testing dataset with a ratio of 4:1. Model optimization was conducted on the training set through 5-fold cross-validation, and we fine-tuned hyperparameters using Bayesian optimization. To ensure the reproducibility of our experiments, we fixed the random state seed across all model runs.

To assess the effectiveness of our models comprehensively, we employed a battery of evaluation metrics, including AUC, sensitivity, specificity, positive predictive value (PPV), negative predictive value (NPV), and F1 score. To mitigate the risk of overfitting and to derive robust confidence intervals (CIs), we implemented a bootstrapping strategy. This involved conducting 100 experiments by randomly resampling the training and testing datasets. In addition to traditional performance metrics, we delved into the interpretability of the XGBoost models. Specifically, we computed SHAP values ^20^ to gain insights into the inner workings of the ML algorithms and to identify the core contribution predictors. This approach aimed to unveil the high-risk factors associated with EOCRC, shedding light on the most influential features in our prediction model.

## 3. Results

Table 1 provides an overview of the identified study cohorts after PSM for both CC and RC across various prediction windows. Notably, CC cases outnumber RC cases, with approximately twice as many CC cases. Patients in the RC groups were slightly older compared to those in the CC group. Sex distribution in the RC groups was closer to parity (2:3 male to female) than in the CC group (2:5 male to female). Both RC and CC groups exhibited diverse racial and ethnic representation. In addition, as the prediction window lengthened, the number of cases decreased. Specifically, there were 560 (0-year), 560 (1-year), 383 (3-year), and 225 (5-year) RC cases, and 1358 (0-year), 1358 (1-year), 884 (3-year), and 532 (5-year) CC cases in each prediction window.

**Table 1.** Descriptive statistics in case and control groups.

| Baseline variables | CC cases<br>(n = 1358) | CC controls<br>(n = 6790) | RC cases<br>(n= 560) | RC controls<br>(n= 2800) |
| --- | --- | --- | --- | --- |
| Age, mean (std) | 36.54 (5.88) | 36.69 (5.73) | 37.70 (5.70) | 36.80 (5.53) |
| Sex, N (%) |  |  |  |  |
| Female | 938 (69.07) | 4461 (65.70) | 323 (57.68) | 1617 (57.75) |
| Male | 420 (30.93) | 2329 (34.30) | 237 (42.32) | 1183 (42.25) |
| Race and Ethnicity, N (%) |  |  |  |  |
| Hispanic | 338 (24.89) | 1527 (22.49) | 101 (18.04) | 514 (18.36) |
| Non-Hispanic White | 554 (40.80) | 2893 (42.61) | 239(42.68) | 1212 (43.29) |
| Non-Hispanic Black | 353 (25.99) | 1857 (27.35) | 178 (31.79) | 887 (31.68) |
| Other | 14 (1.03) | 66 (0.97) | 4 (0.71) | 9 (0.32) |
| Unknown | 99 (7.29) | 447 (6.58) | 38 (6.79) | 178 (6.36) |

CC: colon cancer; RC: rectal cancer

Table 2 presents the results of CC prediction using two feature engineering strategies: one excluding CRC-related features and the other excluding cancer-related features. Additional evaluation metrics for CC prediction across all settings can be found in the Supplementary Material (refer to Tables S1-S2). In most cases, tree-based models (XGBoost and RF) outperformed linear models (SVM and LR), yielding higher AUC values. Specifically, after removing CRC-related features, the RF model achieved the highest AUC [95% CI] for the 0-year prediction (0.811 [0.808, 0.814]), while RF performed best for the 1-year (0.748 [0.745, 0.751]), 3-year (0.689 [0.684, 694]), and 5-year (0.686 [0.68, 0.692]) predictions for CC. However, after removing features associated with prior cancers, the model performance decreased: LR achieved AUC [95% CI] values of 0.788 [0.786, 0.791] for 0-year prediction; RF achieved AUC [95% CI] values of 0.716 [0.713, 0.719] for 1-year, 0.684 [0.679, 0.688] for 3-year, and 0.663 [0.658, 0.668] for 5-year prediction. Performance metrics, including specificity, sensitivity, PPV, NPV, and F1 score, exhibited similar trends.

**Table 2.**
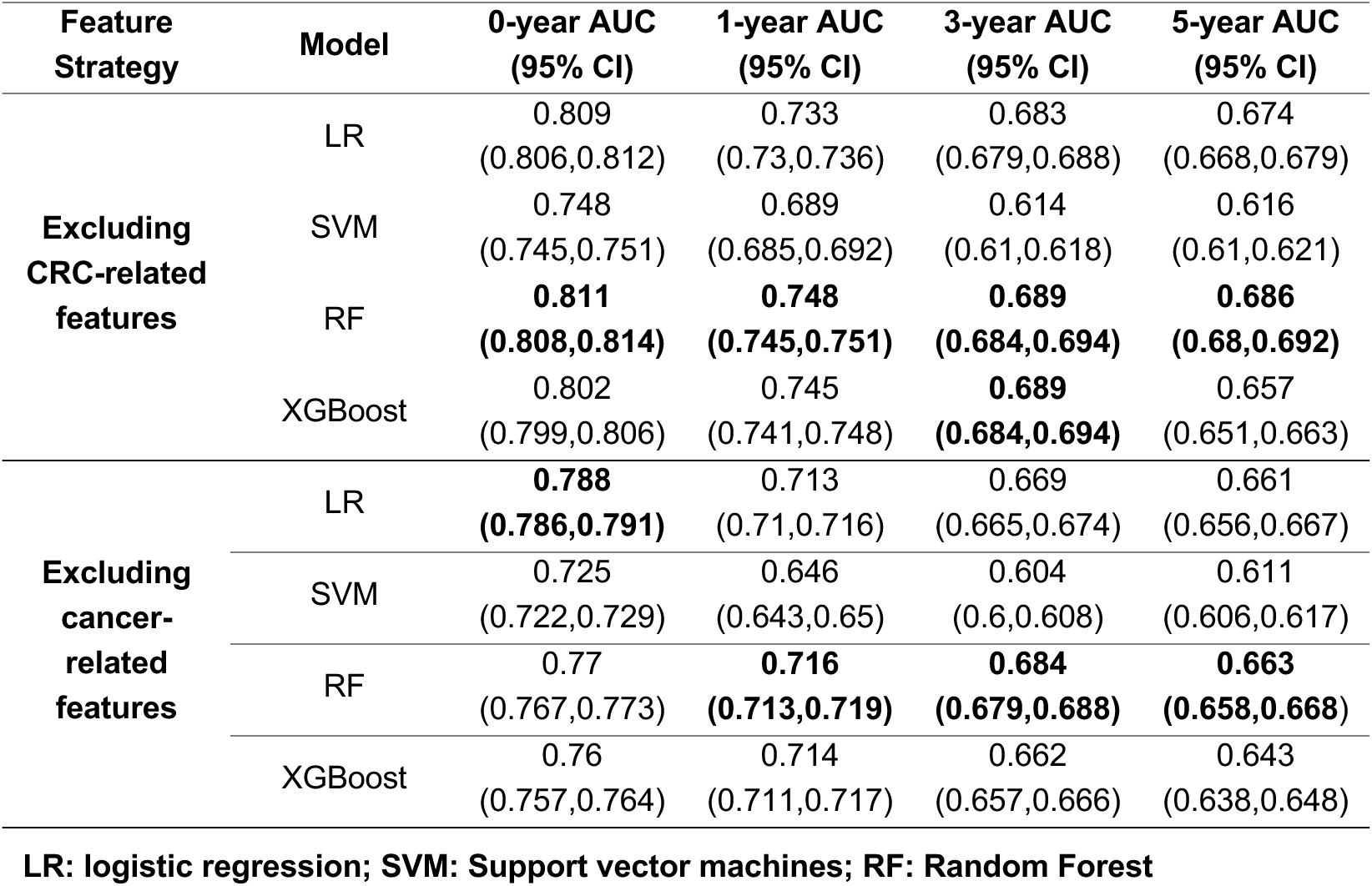
AUC comparison for CC prediction using ML models across different prediction windows (0, 1, 3, and 5 years).

Table 3 provides RC prediction results using the same feature engineering strategies and four prediction windows. Additional evaluation metrics for RC prediction across all settings can be found in the Supplementary Material (refer to Tables S3-S4). Again, after removing CRC-related features, the XGBoost model achieved the highest AUC [95% CI] for the 0-year prediction (0.829 [0.825, 0.834]), while RF performed best for the 1-year (0.771 [0.766,0.777]), and XGBoost did best for 3-year (0.727 [0.721, 0.732]), and 5-year (0.721 [0.713,0.729]) predictions for RC. Eliminating cancer-related features resulted in a performance decrease: XGBoost achieved AUC [95% CI] values of 0.811 [0.806, 0.815] for 0-year prediction. RF achieved AUC [95% CI] values of 0.756 [0.751, 0.76] for 1-year, 0.724 [0.718, 0.73] for 3-year, and 0.711 [0.704, 0.719] for 5-year prediction. Performance metrics exhibited consistent trends.

**Table 3.**
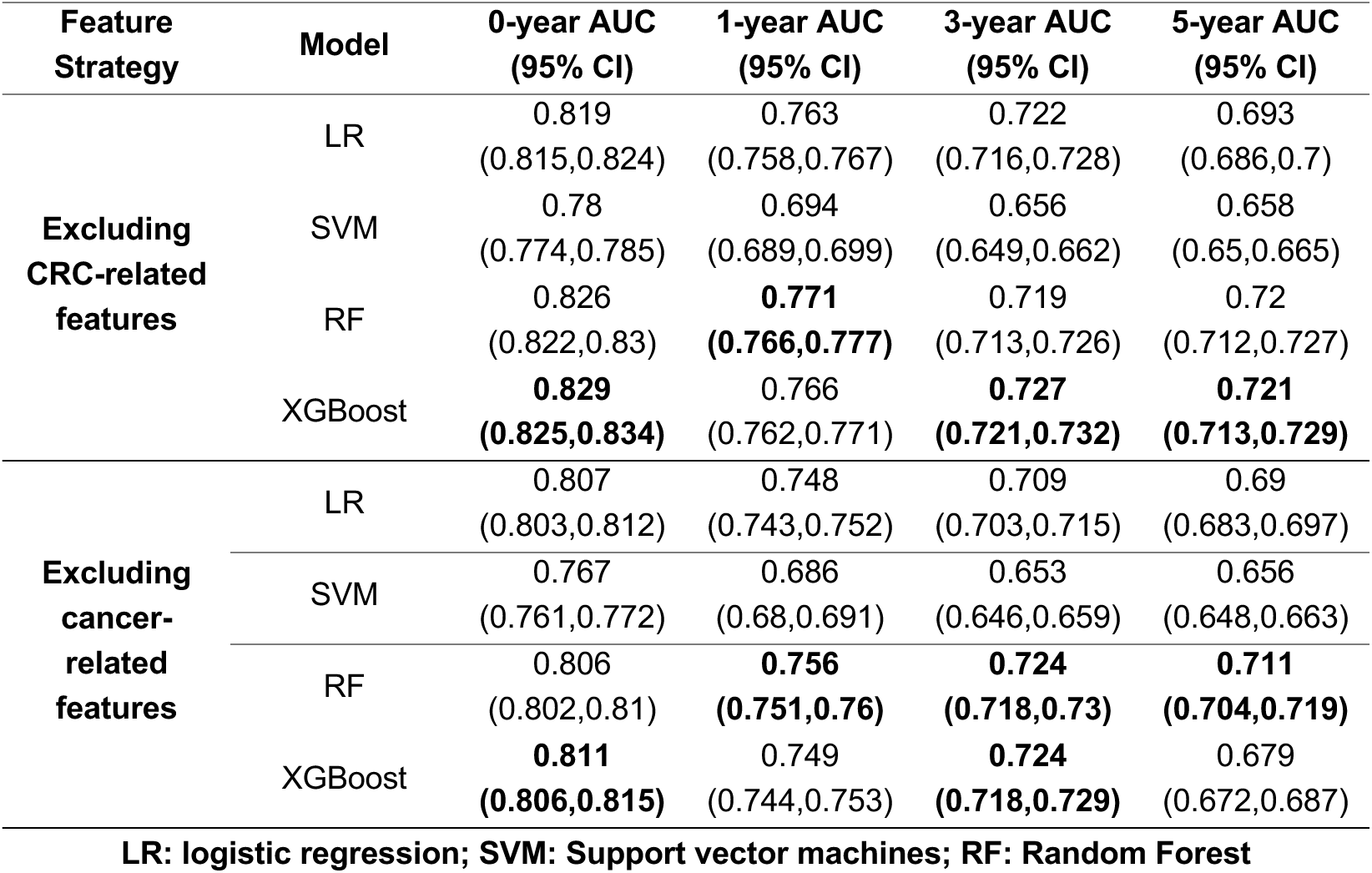
AUC comparison for RC prediction using ML models across different prediction windows (0, 1, 3, and 5 years).

In both the CC and RC prediction tasks, we observed a decline in model performance as the prediction window length increased. Notably, when we removed cancer-related features, the AUC declined. This highlights the pivotal role these features play in enhancing prediction performance. To gain deeper insights into the risk factors associated with these findings, we present SHAP summary plots for CC and RC predictions using two feature engineering strategies and for 0-year and 3-year prediction windows in Figures 3 and 4. Supplementary SHAP summary plots for all other models can be found in the Supplementary Material (Figures S1–S2). Within the CC group, several predictors emerged as positively associated with the risk of CC. Notably, several diagnoses involving various tumors, such as suspected cancer, secondary malignant neoplasm, benign neoplasm of uterus, benign neoplasm of skin, neoplasm of uncertain behavior, neoplasm of uncertain behavior of skin, cancer of other female genital organs and myeloproliferative diseases were identified as influential factors. Gastrointestinal symptoms, encompassing conditions like gastrointestinal hemorrhage, other disorders of intestine, other symptoms involving the abdomen and pelvis, noninfectious gastroenteritis, appendiceal conditions, diverticulosis and diverticulitis, intestinal obstruction without hernia, and disorders of the intestine also exhibited a positive association with CC risk. Additionally, medical procedures related to gastrointestinal diseases and symptoms, including upper gastrointestinal endoscopy, were significantly associated with the development of CC. In the RC group, similar positive predictors were identified, mirroring the trends observed in the CC group, including gastrointestinal symptoms (e.g., gastrointestinal hemorrhage, anal and rectal conditions) and the presence of other cancers or tumors (e.g., secondary malignant neoplasms, benign neoplasms of the uterus or skin). Additionally, the presence of autoimmunity, diseases associated with a potentially weakened immune system (e.g HIV, viral warts and HPV), and conditions like hemorrhoids were linked to a heightened long-term risk of RC. Being underweight was a significant symptom associated with both CC and RC. Conversely, obesity, overweight and normal weight appeared to be negatively associated with RC development. Importantly, after removing cancer-related features from consideration, the significance of anemias surged to the forefront in both the CC and RC groups. These included indicators such as iron deficiency anemias and other anemias. Nevertheless, gastrointestinal diseases and immunodeficiency pathological changes remained substantial factors contributing to CC risk, while factors such as HPV and weight retained their significance as primary determinants of RC. The use of anti-inflammatory or antirheumatic medications were associated with decreased risk of RC.

**Figure 3.**
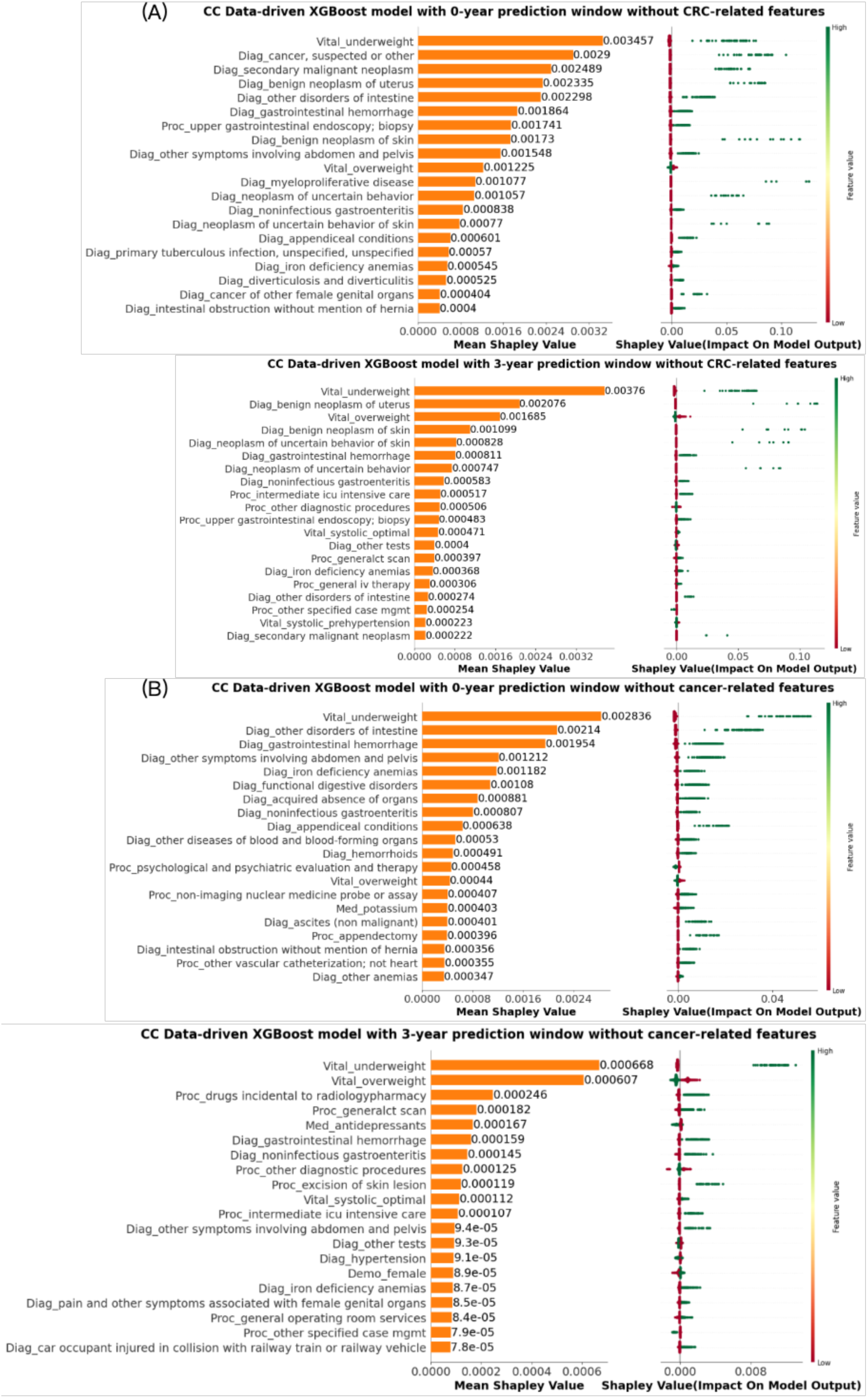
SHAP summary plot of the top 20 features in CC prediction using XGBoost models with 0-year and 3-year prediction windows: (A) excluding CRC-related features; (B) excluding cancer-related features. The prefix before the “_” in the y-axis labels of plots indicates the source of the corresponding features in the PCORnet data model. Specifically, these sources are: Diagnosis (Diag), Procedure (Proc), Medication (Med), Vital Signs (Vital), and Demographics (Demo).

**Figure 4.**
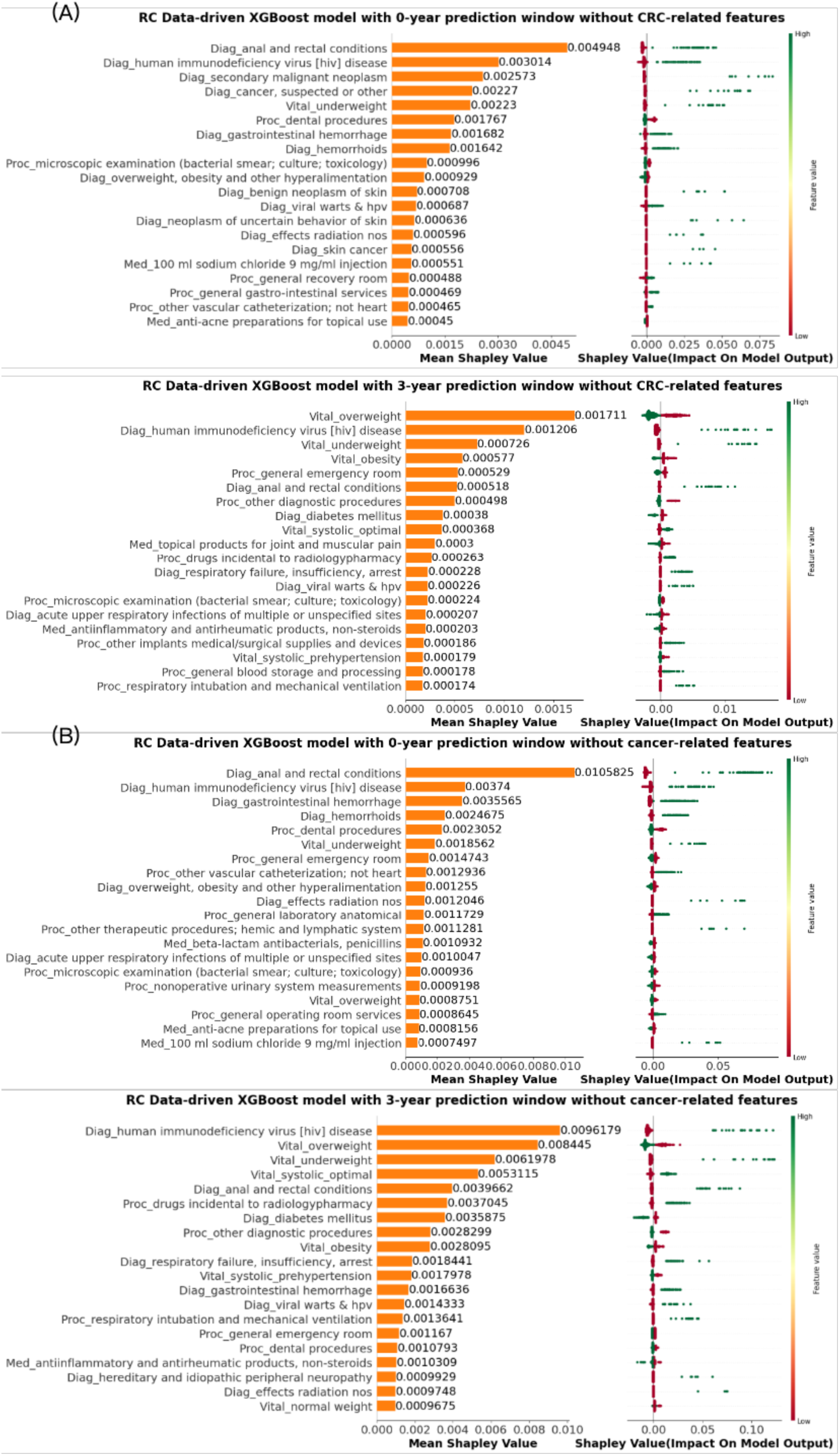
SHAP summary plot of the top 20 features in RC prediction using XGBoost models with 0-year and 3-year prediction windows: (A) excluding CRC-related features; (B) excluding cancer-related features.

## 3. Discussion

In this study, we employed four traditional ML algorithms (i.e., XGBoost, RF, SVM, and LR) and obtained informative results predicting EOCRC using structured EHR data. In most cases, the tree-based models, (XGBoost and RF) outperformed linear models, achieving the best AUC scores for various prediction windows. Additionally, even after excluding cancer diagnosis variables (e.g., pancreatic, skin, thyroid cancer), undergoing cancer-related procedures (e.g., liver biopsy, bone marrow biopsy), cancer treatments (e.g., cisplatin, doxycycline), our models continued to achieve acceptable AUC scores. Immune and digestive system disorders, blood diseases, and secondary cancers were identified as significant predictors.

Most of our experimental findings were consistent with existing published research. Cancer-related diseases and diagnoses emerged as risk factors leading to the diagnosis of EOCRC, both for colon and rectal cancers. For example, uterine cancer was identified as a driver of EOCRC, suggesting a potential genetic association between these malignancies in younger patients.^31^ Research also demonstrates that the incidence rate of second primary cancers among survivors is significantly higher than cancer in the general population, and survivors experience notable morbidity and mortality from their cancer treatment.^32^ Additionally, the use of CT scans for other medical reasons could contribute to the incidental identification of EOCRC cases.^33^ Notably, we know that some forms of cancer treatment (e.g., radiation) predisposes one to an increased risk for secondary malignancies, including EOCRC, particularly in patients surviving a childhood cancer.^34^

Inflammatory bowel diseases (IBD) are well established risk factors for colorectal cancers, particularly during young adulthood. The chronic inflammation associated with IBD leads to the release of growth cytokines, excess blood flow, and metabolic free radicals, all of which contribute to the heightened risk of developing colorectal cancer.^35^ Therapies for IBD sometimes involve immune suppression, another known risk factor for cancers. Furthermore, many gastrointestinal diseases can cause malabsorption or malnutrition,^36^ resulting in patients being underweight which can also contribute to immune dysfunction or suppression.^37^ However, overweight patients were at low risk of EOCRC as our analysis demonstrated despite emerging evidence that being overweight may be associated with an increased risk of tumor recurrence and colorectal carcinogenesis.^38,39^ The temporal use of antibiotics in relation to subsequent development of EOCRC is an interesting finding as it supports several previously reported roles that the gut microbiome may plan in colorectal cancer protection and development.^40^ Our analysis highlighted that the diagnosis of iron deficiency anemia pre-dated colon cancer, but had less association with rectal cancers. It is logical, given that colon cancers are situated more proximal in the gastrointestinal tract, causing occult chronic blood loss and subsequent anemia rather than overt gross bleeding as is typically evident from rectal cancers.

Additionally, our study observed a significantly higher incidence of colorectal cancer cases among HIV-infected patients compared to HIV-uninfected individuals.^41^ The heightened risk can be attributed to disruptions in immune function caused by immunodeficiency, which exposes individuals to a higher susceptibility against cancer-causing viruses, including HPV, EBV, KSHV, etc., as evidenced in our analysis. ^42^ Another notable finding was the association between colon cancer and diseases of myeloproliferative disease. Similar to other cancers, the potential link could be related to genetics, treatments that induce DNA damage that could predispose to EOCRC, and chronic immune dysregulation. Overall, our study sheds light on the complex interplay between inflammatory bowel diseases, malnutrition, immune function, and specific blood-related diseases in the development of CRC. Understanding these relationships is crucial in advancing our knowledge of EOCRC risk factors and devising targeted interventions for at-risk populations.

Our study does have several limitations. First, the mechanism through which identified medical factors are associated with EOCRC is speculative. For example, CT scans contributed significantly to the model’s performance, but the specific reasons are unclear. EHRs didn’t record the reason why patients underwent CT scans. Perhaps some patients obtained CT scans because of symptoms related to undiagnosed CRC while others received CT scans for other reasons with the incidental finding of CRC. It is less likely that CT scans could be associated with causing CRC due to radiation exposure. For that to occur, the cumulative lifetime exposure would need to be very high with exposure over a number of decades for that to occur. Perhaps CT imaging itself is just a surrogate for access to care whereby EOCRC is more likely to be eventually diagnosed as opposed to patients who might expire for other reasons with CRC, but prior to a diagnosis. Second, the exclusion of confounder samples and features posed difficulties, given the lack of universally accepted standards for phenotype definitions and ambiguous descriptions. These challenges hindered the design of the most optimal experiment.^43^ Third, our experiments are carried out based on the EHR data, which inherently contains flaws, including missing values and potential mistakes in records. Efforts were made to fill missing values, but comprehensive amendments remained challenging. The characteristics of the EHR data, such as temporality, irregularity, sparsity, and data imbalance, can result in abnormal outcomes when applying machine learning models.^44,45^ Moreover, systematic bias, such as erroneous use of ICD codes due to strategic billing, may impact data-driven predictions.^46^ The EHR data utilized in the OneFlorida+ dataset are overwhelmingly hospital-based data, which may further introduce selection bias in that ambulatory practices (where most relatively healthy patients receive their routine care) is inconsistently represented. Despite these limitations, we believe our model provides interesting insights into medical variables that pre-date and are associated with EOCRC.

## 4. Conclusion

In conclusion, our study demonstrated the potential of traditional ML algorithms in predicting EOCRC using real-world data for individuals below the screening age guideline. The identification of significant predictors and their consistency with academic research findings provide valuable insights for pursuing additional hypotheses or targeting potential patients at risk for EOCRC. However, addressing the challenges and limitations related to data quality, experimental design, and ML models’ development is essential for improving the accuracy and reliability of EOCRC prediction models. Future research should focus on refining the experimental design, exploring alternative feature selection techniques, incorporating LLM based on both ambulatory and inpatient data, and integrating domain knowledge to enhance the performance of the prediction models. Ultimately, these efforts will contribute to early detection and better management of CRC, with the goal to improve patient outcomes.

## Data Availability

All data produced in the present study are available upon reasonable request to the authors

## Notes

### Competing Interest Statement

The authors have declared no competing interest.

### Funding Statement

This study was funded by the UF CTSI Precision Health Initiative.

### Author Declarations

The study has been approved and the requirement to obtain any informed consent has been waived by the University of Florida Institutional Review Board (protocol no. IRB202201561). The research does not involve greater than minimal risk for participation. Analyses only involve the secondary analysis of data that are either limited data sets or de-identified. Our research team has no direct contact with human subjects. All methods were carried out in accordance with relevant guidelines and regulations.

