## Supplementary Material for "Predicting Early-Onset Colorectal Cancer in Individuals Below Screening Age Using Machine Learning and Real-World Data"

**Table S1.** The performance of CC prediction using ML models across different prediction windows (0, 1, 3, and 5 years), excluding CRC-related features.

| Prediction Window | Model | AUC (95% CI) | Sensitivity (95% CI) | Specificity (95% CI) | PPV (95% CI) | NPV (95% CI) | F1 (95% CI) |
|---|---|---|---|---|---|---|---|
| 0-year | LR | 0.809 (0.806,0.812) | 0.674 (0.663,0.685) | 0.807 (0.796,0.818) | 0.421 (0.41,0.433) | 0.926 (0.924,0.928) | **0.849 (0.847,0.851)** |
| | SVM | 0.748 (0.745,0.751) | 0.633 (0.62,0.647) | 0.754 (0.742,0.767) | 0.347 (0.338,0.356) | 0.912 (0.91,0.914) | 0.822 (0.82,0.824) |
| | RF | **0.811 (0.808,0.814)** | **0.686 (0.673,0.698)** | 0.791 (0.779,0.804) | 0.407 (0.396,0.418) | **0.927 (0.925,0.929)** | 0.78 (0.778,0.783) |
| | XGBoost | 0.802 (0.799,0.806) | 0.61 (0.601,0.619) | **0.887 (0.879,0.895)** | **0.533 (0.518,0.548)** | 0.92 (0.918,0.921) | 0.758 (0.755,0.76) |
| 1-year | LR | 0.733 (0.73,0.736) | 0.595 (0.581,0.609) | 0.758 (0.745,0.772) | 0.338 (0.33,0.347) | **0.904 (0.902,0.906)** | **0.816 (0.814,0.819)** |
| | SVM | 0.689 (0.685,0.692) | 0.534 (0.514,0.554) | 0.763 (0.745,0.782) | 0.329 (0.317,0.342) | 0.893 (0.89,0.895) | 0.797 (0.795,0.799) |
| | RF | **0.748 (0.745,0.751)** | **0.613 (0.597,0.63)** | 0.758 (0.742,0.774) | 0.351 (0.338,0.363) | 0.909 (0.906,0.911) | 0.763 (0.76,0.765) |
| | XGBoost | 0.745 (0.741,0.748) | 0.561 (0.546,0.576) | **0.816 (0.801,0.831)** | **0.401 (0.384,0.417)** | **0.904 (0.902,0.906)** | 0.758 (0.755,0.76) |
| 3-year | LR | 0.683 (0.679,0.688) | **0.574 (0.554,0.593)** | 0.713 (0.695,0.731) | 0.296 (0.288,0.305) | **0.894 (0.891,0.897)** | **0.789 (0.786,0.792)** |
| | SVM | 0.614 (0.61,0.618) | 0.487 (0.466,0.508) | 0.709 (0.688,0.73) | 0.262 (0.254,0.27) | 0.874 (0.871,0.876) | 0.767 (0.765,0.77) |
| | RF | **0.689 (0.684,0.694)** | 0.42 (0.405,0.435) | **0.885 (0.872,0.898)** | **0.45 (0.433,0.467)** | 0.884 (0.882,0.886) | 0.763 (0.76,0.766) |
| | XGBoost | **0.689 (0.684,0.694)** | 0.455 (0.434,0.475) | 0.845 (0.825,0.864) | 0.414 (0.393,0.435) | 0.886 (0.884,0.888) | 0.757 (0.754,0.759) |
| 5-year | LR | 0.674 (0.668,0.679) | **0.59 (0.565,0.616)** | 0.688 (0.664,0.712) | 0.29 (0.28,0.3) | **0.895 (0.892,0.899)** | **0.793 (0.789,0.796)** |
| | SVM | 0.616 (0.61,0.621) | 0.57 (0.542,0.597) | 0.637 (0.61,0.664) | 0.251 (0.242,0.259) | 0.883 (0.879,0.887) | 0.767 (0.764,0.77) |
| | RF | **0.686 (0.68,0.692)** | 0.552 (0.528,0.575) | 0.753 (0.732,0.775) | 0.333 (0.317,0.348) | **0.895 (0.892,0.898)** | 0.758 (0.754,0.761) |
| | XGBoost | 0.657 (0.651,0.663) | 0.406 (0.388,0.425) | **0.874 (0.858,0.89)** | **0.436 (0.413,0.459)** | 0.881 (0.878,0.883) | 0.757 (0.754,0.76) |

**Table S2.** The performance of CC prediction using ML models across different prediction windows (0, 1, 3, and 5 years), excluding cancer-related features.

| Prediction Window | Model | AUC (95% CI) | Sensitivity (95% CI) | Specificity (95% CI) | PPV (95% CI) | NPV (95% CI) | F1 (95% CI) |
|---|---|---|---|---|---|---|---|
| 0-year | LR | **0.788 (0.786,0.791)** | 0.672 (0.661,0.683) | 0.777 (0.766,0.787) | 0.382 (0.373,0.391) | **0.923 (0.921,0.925)** | **0.829 (0.827,0.831)** |
| | SVM | 0.725 (0.722,0.729) | 0.63 (0.616,0.645) | 0.725 (0.712,0.738) | 0.32 (0.312,0.327) | 0.908 (0.906,0.911) | 0.809 (0.807,0.811) |
| | RF | 0.77 (0.767,0.773) | **0.7 (0.684,0.715)** | 0.71 (0.695,0.726) | 0.333 (0.325,0.342) | **0.923 (0.921,0.926)** | 0.758 (0.756,0.761) |
| | XGBoost | 0.76 (0.757,0.764) | 0.595 (0.583,0.607) | **0.82 (0.809,0.831)** | **0.407 (0.397,0.417)** | 0.911 (0.909,0.913) | 0.758 (0.755,0.76) |
| 1-year | LR | 0.713 (0.71,0.716) | 0.601 (0.587,0.616) | **0.721 (0.707,0.736)** | **0.307 (0.301,0.314)** | 0.901 (0.899,0.903) | **0.804 (0.801,0.806)** |
| | SVM | 0.646 (0.643,0.65) | 0.549 (0.533,0.566) | 0.686 (0.67,0.702) | 0.263 (0.258,0.269) | 0.885 (0.882,0.887) | 0.777 (0.775,0.779) |
| | RF | **0.716 (0.713,0.719)** | **0.654 (0.639,0.668)** | 0.671 (0.657,0.685) | 0.289 (0.282,0.296) | **0.907 (0.905,0.91)** | 0.758 (0.755,0.76) |
| | XGBoost | 0.714 (0.711,0.717) | 0.614 (0.599,0.629) | 0.707 (0.691,0.722) | 0.301 (0.294,0.309) | 0.902 (0.9,0.905) | 0.758 (0.755,0.76) |
| 3-year | LR | 0.669 (0.665,0.674) | 0.575 (0.554,0.596) | 0.69 (0.67,0.71) | 0.281 (0.273,0.29) | 0.891 (0.888,0.894) | **0.783 (0.78,0.786)** |
| | SVM | 0.604 (0.6,0.608) | 0.523 (0.496,0.55) | 0.656 (0.629,0.683) | 0.246 (0.238,0.254) | 0.874 (0.871,0.877) | 0.773 (0.771,0.775) |
| | RF | **0.684 (0.679,0.688)** | **0.587 (0.565,0.61)** | 0.692 (0.671,0.714) | 0.29 (0.279,0.3) | **0.895 (0.892,0.898)** | 0.764 (0.761,0.767) |
| | XGBoost | 0.662 (0.657,0.666) | 0.494 (0.469,0.518) | **0.758 (0.735,0.781)** | **0.313 (0.3,0.325)** | 0.883 (0.88,0.886) | 0.757 (0.754,0.759) |
| 5-year | LR | 0.661 (0.656,0.667) | **0.606 (0.581,0.63)** | 0.656 (0.631,0.68) | 0.272 (0.263,0.282) | **0.895 (0.891,0.898)** | **0.777 (0.773,0.78)** |
| | SVM | 0.611 (0.606,0.617) | 0.597 (0.566,0.629) | 0.6 (0.568,0.632) | 0.242 (0.234,0.251) | 0.885 (0.881,0.889) | 0.766 (0.762,0.769) |
| | RF | **0.663 (0.658,0.668)** | 0.596 (0.569,0.623) | 0.668 (0.643,0.693) | 0.281 (0.269,0.294) | 0.894 (0.891,0.898) | 0.757 (0.754,0.76) |
| | XGBoost | 0.643 (0.638,0.648) | 0.532 (0.505,0.558) | **0.706 (0.68,0.731)** | **0.283 (0.272,0.293)** | 0.885 (0.881,0.888) | 0.757 (0.754,0.76) |

**Table S3.** The performance of RC prediction using ML models across different prediction windows (0, 1, 3, and 5 years), excluding CRC-related.

| Prediction Window | Model | AUC (95% CI) | Sensitivity (95% CI) | Specificity (95% CI) | PPV (95% CI) | NPV (95% CI) | F1 (95% CI) |
|---|---|---|---|---|---|---|---|
| **0-year** | LR | 0.819 (0.815,0.824) | 0.688 (0.674,0.701) | 0.839 (0.827,0.851) | **0.48 (0.462,0.498)** | 0.932 (0.929,0.934) | **0.858 (0.855,0.861)** |
| | SVM | 0.78 (0.774,0.785) | 0.684 (0.672,0.696) | 0.761 (0.746,0.775) | 0.379 (0.366,0.392) | 0.927 (0.924,0.93) | 0.838 (0.835,0.841) |
| | RF | 0.826 (0.822,0.83) | **0.7 (0.688,0.713)** | 0.826 (0.812,0.841) | 0.469 (0.452,0.487) | **0.934 (0.932,0.937)** | 0.775 (0.771,0.778) |
| | XGBoost | **0.829 (0.825,0.834)** | 0.666 (0.656,0.676) | **0.877 (0.865,0.888)** | 0.54 (0.523,0.558) | 0.931 (0.929,0.933) | 0.758 (0.755,0.761) |
| **1-year** | LR | 0.763 (0.758,0.767) | **0.635 (0.619,0.65)** | 0.787 (0.773,0.802) | **0.458 (0.446,0.471)** | 0.894 (0.891,0.897) | 0.788 (0.784,0.792) |
| | SVM | 0.694 (0.689,0.699) | 0.609 (0.592,0.625) | 0.705 (0.689,0.72) | 0.301 (0.293,0.308) | 0.903 (0.9,0.906) | **0.797 (0.794,0.8)** |
| | RF | **0.771 (0.766,0.777)** | 0.583 (0.564,0.603) | 0.803 (0.784,0.823) | 0.424 (0.403,0.444) | **0.918 (0.915,0.92)** | 0.77 (0.767,0.773) |
| | XGBoost | 0.766 (0.762,0.771) | 0.557 (0.539,0.574) | **0.818 (0.802,0.834)** | 0.432 (0.413,0.451) | 0.916 (0.914,0.918) | 0.758 (0.755,0.761) |
| **3-year** | LR | 0.722 (0.716,0.728) | **0.598 (0.577,0.618)** | 0.72 (0.704,0.736) | 0.382 (0.371,0.393) | 0.893 (0.888,0.897) | 0.764 (0.759,0.768) |
| | SVM | 0.656 (0.649,0.662) | 0.545 (0.522,0.567) | 0.679 (0.654,0.705) | 0.286 (0.274,0.298) | 0.896 (0.892,0.9) | **0.786 (0.782,0.789)** |
| | RF | 0.719 (0.713,0.726) | 0.525 (0.507,0.542) | 0.812 (0.794,0.83) | 0.394 (0.377,0.411) | **0.901 (0.898,0.904)** | 0.758 (0.754,0.762) |
| | XGBoost | **0.727 (0.721,0.732)** | 0.512 (0.495,0.528) | **0.858 (0.839,0.877)** | **0.469 (0.445,0.492)** | **0.901 (0.898,0.904)** | 0.758 (0.754,0.762) |
| **5-year** | LR | 0.693 (0.686,0.7) | 0.616 (0.586,0.645) | 0.668 (0.64,0.697) | 0.353 (0.336,0.37) | 0.898 (0.892,0.903) | 0.767 (0.761,0.774) |
| | SVM | 0.658 (0.65,0.665) | **0.648 (0.617,0.679)** | 0.647 (0.62,0.674) | 0.285 (0.274,0.295) | 0.904 (0.899,0.91) | **0.781 (0.776,0.786)** |
| | RF | 0.72 (0.712,0.727) | 0.526 (0.503,0.548) | **0.766 (0.736,0.796)** | **0.394 (0.369,0.418)** | 0.91 (0.904,0.915) | 0.759 (0.754,0.764) |
| | XGBoost | **0.721 (0.713,0.729)** | 0.473 (0.452,0.494) | 0.72 (0.697,0.743) | 0.343 (0.328,0.359) | **0.915 (0.911,0.919)** | 0.756 (0.751,0.761) |

**Table S4.** The performance of RC prediction using ML models across different prediction windows (0, 1, 3, and 5 years), excluding cancer-related features.

| Prediction Window | Model | AUC (95% CI) | Sensitivity (95% CI) | Specificity (95% CI) | PPV (95% CI) | NPV (95% CI) | F1 (95% CI) |
|---|---|---|---|---|---|---|---|
| **0-year** | LR | 0.807 (0.803,0.812) | 0.687 (0.672,0.701) | **0.821 (0.809,0.834)** | **0.45 (0.435,0.465)** | 0.93 (0.928,0.932) | **0.848 (0.844,0.851)** |
| | SVM | 0.767 (0.761,0.772) | 0.684 (0.669,0.7) | 0.752 (0.738,0.767) | 0.366 (0.355,0.377) | 0.924 (0.921,0.926) | 0.829 (0.826,0.832) |
| | RF | 0.806 (0.802,0.81) | **0.714 (0.701,0.727)** | 0.783 (0.77,0.795) | 0.408 (0.395,0.421) | **0.933 (0.93,0.935)** | 0.759 (0.756,0.762) |
| | XGBoost | **0.811 (0.806,0.815)** | 0.701 (0.687,0.714) | 0.817 (0.805,0.83) | 0.448 (0.434,0.462) | **0.933 (0.93,0.935)** | 0.758 (0.755,0.761) |
| **1-year** | LR | 0.748 (0.743,0.752) | 0.639 (0.623,0.654) | 0.769 (0.756,0.782) | 0.366 (0.354,0.378) | 0.915 (0.912,0.918) | **0.804 (0.801,0.808)** |
| | SVM | 0.686 (0.68,0.691) | 0.602 (0.583,0.621) | 0.713 (0.695,0.73) | 0.303 (0.295,0.312) | 0.901 (0.898,0.904) | 0.793 (0.791,0.796) |
| | RF | **0.756 (0.751,0.76)** | **0.642 (0.627,0.656)** | **0.778 (0.765,0.791)** | 0.376 (0.365,0.387) | **0.917 (0.914,0.919)** | 0.758 (0.755,0.761) |
| | XGBoost | 0.749 (0.744,0.753) | 0.623 (0.606,0.641) | **0.778 (0.761,0.795)** | **0.378 (0.363,0.394)** | 0.913 (0.91,0.916) | 0.758 (0.755,0.761) |
| **3-year** | LR | 0.709 (0.703,0.715) | **0.626 (0.607,0.644)** | 0.723 (0.704,0.741) | 0.323 (0.312,0.335) | 0.907 (0.904,0.91) | **0.789 (0.785,0.793)** |
| | SVM | 0.653 (0.646,0.659) | 0.586 (0.558,0.613) | 0.684 (0.658,0.71) | 0.286 (0.275,0.298) | 0.894 (0.89,0.898) | 0.785 (0.782,0.789) |
| | RF | **0.724 (0.718,0.73)** | 0.567 (0.548,0.586) | **0.804 (0.786,0.821)** | **0.385 (0.371,0.4)** | 0.904 (0.9,0.907) | 0.758 (0.754,0.762) |
| | XGBoost | **0.724 (0.718,0.729)** | 0.607 (0.581,0.633) | 0.756 (0.731,0.782) | 0.369 (0.349,0.389) | **0.908 (0.904,0.912)** | 0.758 (0.754,0.762) |
| **5-year** | LR | 0.69 (0.683,0.697) | 0.631 (0.604,0.659) | 0.703 (0.676,0.73) | 0.326 (0.309,0.343) | 0.907 (0.903,0.911) | **0.795 (0.79,0.801)** |
| | SVM | 0.656 (0.648,0.663) | 0.632 (0.602,0.661) | 0.661 (0.632,0.69) | 0.291 (0.278,0.304) | 0.903 (0.897,0.908) | 0.78 (0.775,0.785) |
| | RF | **0.711 (0.704,0.719)** | **0.672 (0.645,0.699)** | 0.687 (0.66,0.713) | **0.325 (0.31,0.341)** | **0.915 (0.91,0.92)** | 0.756 (0.751,0.762) |
| | XGBoost | 0.679 (0.672,0.687) | 0.61 (0.58,0.64) | **0.706 (0.677,0.734)** | 0.322 (0.306,0.338) | 0.903 (0.898,0.908) | 0.756 (0.751,0.761) |

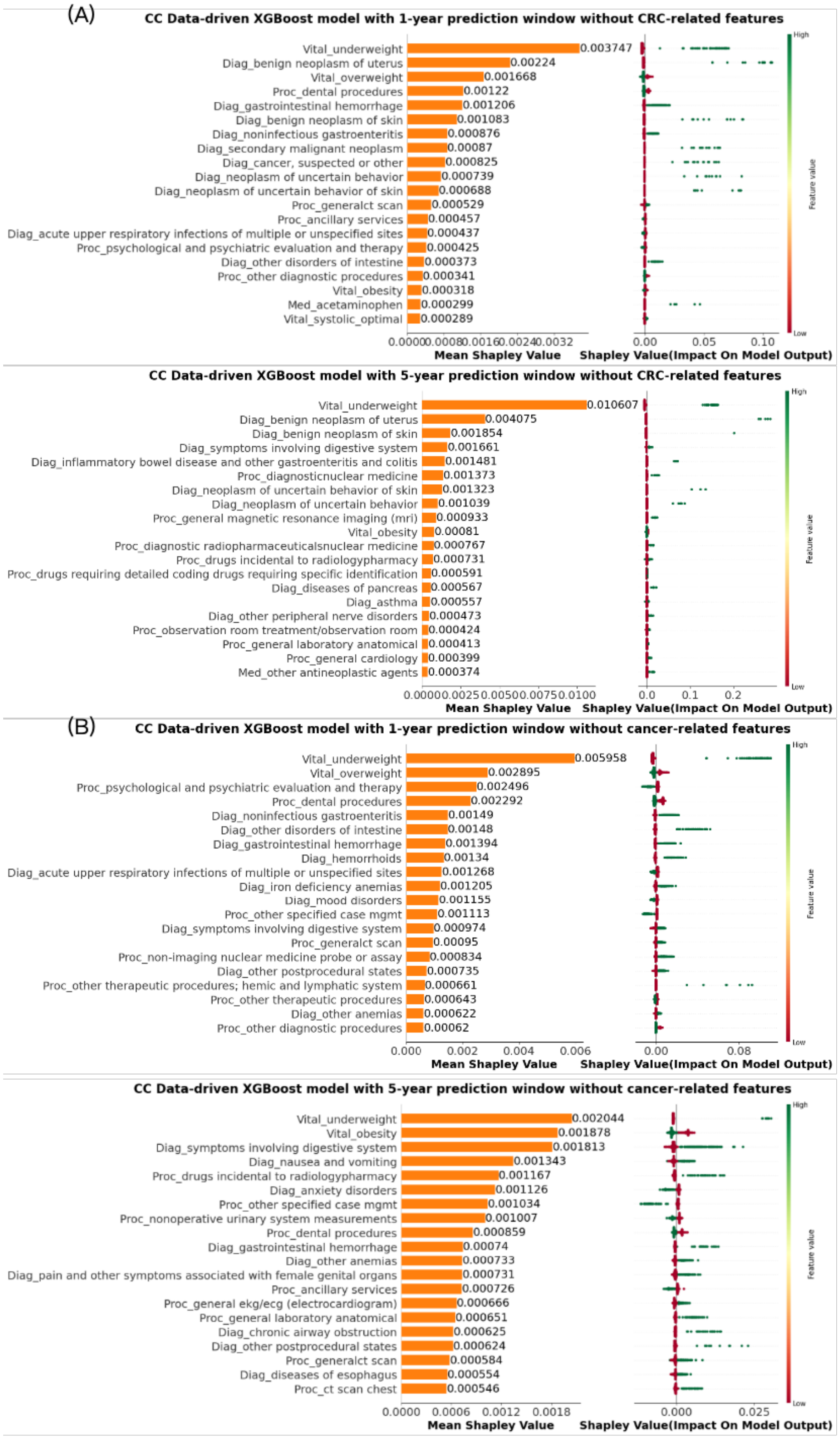


**Figure S1.** SHAP summary plot of the top 20 features in CC prediction using XGBoost models with 1-year and 5-year prediction windows: (A) excluding CRC-related features; (B) excluding cancer-related features. The prefix before the "_" in the y-axis labels of plots indicates the source of the corresponding features in the PCORnet data model. Specifically, these sources are: Diagnosis (Diag), Procedure (Proc), Medication (Med), Vital Signs (Vital), and Demographics (Demo).

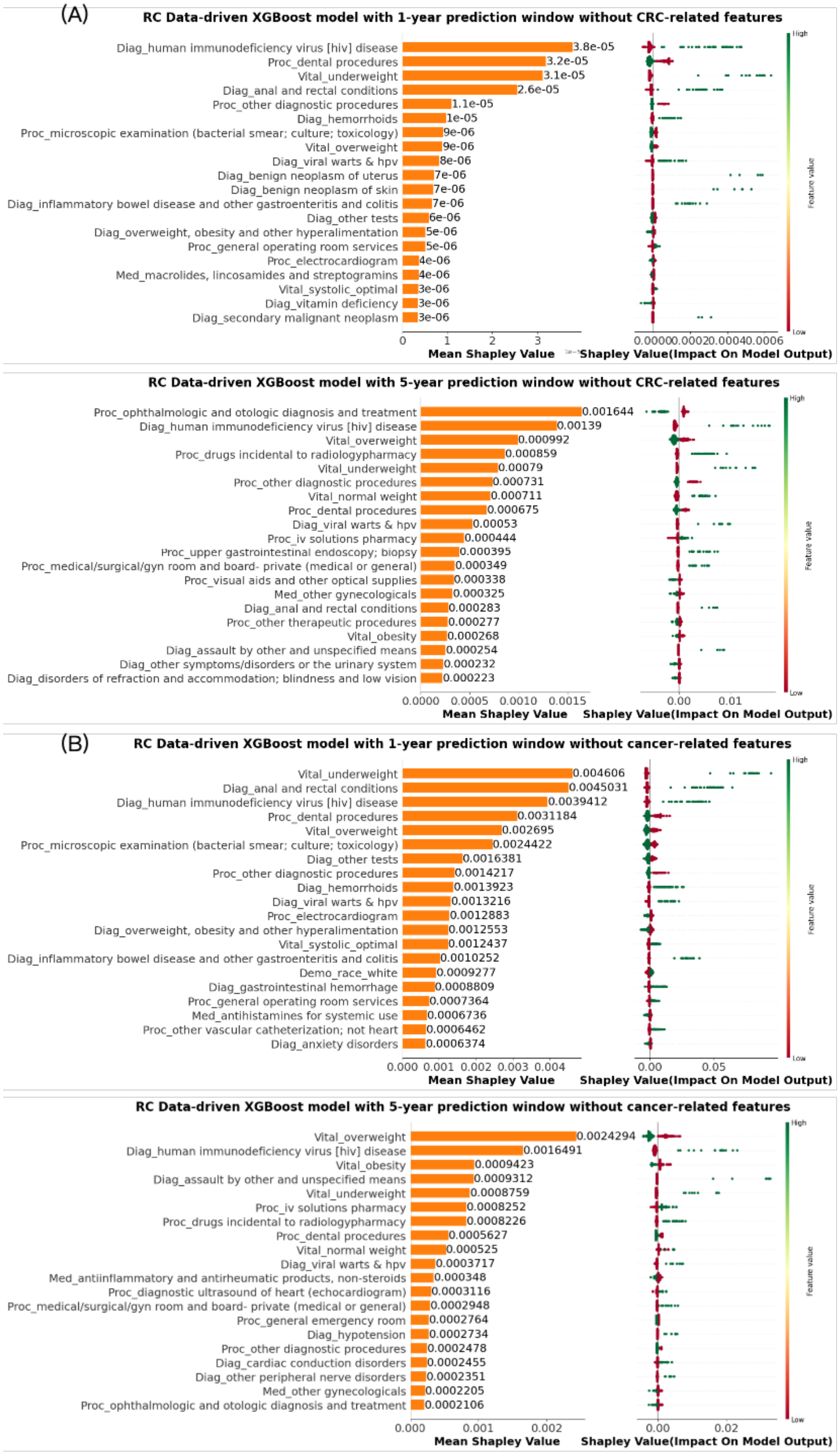


**Figure S2.** SHAP summary plot of the top 20 features in RC prediction using XGBoost models with 1-year and 5-year prediction windows: (A) excluding CRC-related features; (B) excluding cancer-related features.
